# Performance evaluation of an under-mattress sleep sensor versus polysomnography in *≥* 400 nights with healthy and unhealthy sleep

**DOI:** 10.1101/2024.09.09.24312921

**Authors:** Jack Manners, Eva Kemps, Bastien Lechat, Peter Catcheside, Danny Eckert, Hannah Scott

**Author notes:** Co-senior authors (Danny Eckert, Hannah Scott).

## Abstract

Consumer sleep trackers can provide useful insight into sleep and sleep patterns. However, large scale performance evaluation studies against direct sleep measures are needed to comprehensively understand sleep tracker accuracy. This study evaluated performance of an under-mattress sensor to estimate sleep and wake versus polysomnography, during multiple in-laboratory protocols in a large sample including individuals with and without sleep disorders and during day versus night sleep opportunities.

183 participants (51% male, mean[SD] age=45[18] years) attended the sleep laboratory for a research study that included simultaneous polysomnography and under-mattress sensor (Withings Sleep Analyzer [WSA]) recordings. Epoch-by-epoch analyses with confusion matrices were used to determine accuracy, sensitivity, and specificity of the WSA versus polysomnography. Bland-Altman plots examined bias in sleep duration, efficiency, onset-latency, and wake after sleep onset.

Overall WSA sleep-wake classification accuracy was 83%, sensitivity 95%, and specificity 37%. The WSA significantly overestimated total sleep time (48[81]minutes), Sleep efficiency (9[15]%), sleep onset latency (6[26]), and underestimated wake after sleep onset (54[78]), p<0.05. Accuracy and specificity were higher for night versus daytime sleep opportunities in healthy individuals (89% and 47% versus 82% and 26% respectively, p<0.05). Accuracy and sensitivity were also higher for healthy individuals (89% and 97%) versus those with sleep disorders (81% and 91%, p<0.05).

WSA performance is comparable to other consumer sleep trackers, with high sensitivity but poor specificity compared to polysomnography. Poorer accuracy and specificity during daytime versus night-time sleep opportunities is likely due to increased wake time and reduced sleep efficiency. Contactless, under-mattress sleep sensors show promise for accurate sleep monitoring, noting the tendency to over-estimate sleep particularly where wake time is high.

## Introduction

Sleep is critical for optimal daytime function, performance, safety, and health. Thus, accurate and reliable estimates of sleep are important to help manage these key outcomes. However, sleep is difficult to objectively evaluate, particularly over extended multi-day periods. This is partly because gold-standard quantification of sleep, polysomnography, is expensive and complex to administer over multiple nights. Wearable and ”nearable” (devices that are not in direct contact with an individual) sleep tracking devices are used to simplify sleep estimation in the home, such as wrist-based actigraphy, bedside radar, or mattress sensor devices [1, 2]. Deciding which devices are best suited for sleep estimation requires critical appraisal of device performance and practicality. Accordingly, the present study evaluated the performance of an under-mattress device, the Withings Sleep Analyzer (WSA), to evaluate sleep in a diverse cohort and various sleep opportunities (day and night).

To help circumvent cost and measurement complexity for multi-day assessments, wearable sleep trackers, such as actigraphy based devices, are often used to infer wake and sleep from body movements. This approach is sensitive to detect sleep but cannot reliably discriminate wake from sleep when people lie still awake [3]. Additional signals such as heart rate, heart rate variability, and breathing can improve device performance, but poor specificity remains a key limitation of this technology. Another problem is that wearable devices require the user to ensure the device is charged and properly worn. Consequently, data loss is common [4], variable [5], and performance can drastically drop when devices are not worn correctly [6]. ”Nearable” sleep trackers that infer sleep from detected motion, including respiratory and cardiac motion, avoid the need to charge or wear any device. These devices are typically designed to be placed on or under the mattress, or at the bedside, with little subsequent intervention required once properly set up [1, 7]. However, some evidence suggests that these devices may be less accurate than wearable counterparts [8]. Thus, rigorous performance evaluation is essential to determine device reliability, practical benefits, and sleep tracking performance.

The WSA has been previously validated in smaller trials (N=18-118 [49-118 nights]) against polysomnography to estimate sleep and identify breathing disturbances [8–11]. The device uses a pneumatic sensor placed under the mattress to detect air-pressure changes, from which movement, sleep, respiration and heart rate are inferred. The device is attractive for both clinical and research use, as once set up and connected to Wi-Fi and mains power, minimal ongoing user input is required. Indeed, recent large scale, long-term monitoring studies that have used this technology have been able to address key sleep health questions that were not previously possible with polysomnography or existing wearables [11–15]. Furthermore, this technology could markedly reduce data loss compared to wearable sleep trackers that require regular device charging and correct daily wear. The unobtrusive nature of this approach is also potentially amenable to patient/participant monitoring in environments less conducive to wearable devices such as hospitals, nursing homes, and in high-risk on-call workplace settings. Given poor specificity of sleep tracking devices, sleep classification may be less accurate in these environments as sleep is more likely to be impaired, compared to in the home or laboratory. Despite this, few devices have been rigorously evaluated across multi-night sleeps, and fewer devices have been assessed to any degree with non-standard sleep schedules. Device accuracy under these conditions is particularly important if they are to be used for readiness evaluation in shift workers, or to examine clinical outcomes in people with sleep disorders. Therefore, comprehensive performance evaluation of the WSA during sleep opportunities of varied timing and in people with and without sleep disordersd remains important to establish the potential utility of nearable sleep trackers for extended multi-day monitoring in more challenging sleep assessment settings.

This study used comprehensive objective measurements of sleep in several laboratory sleep research studies to evaluate the classification accuracy of the WSA compared to polysomnography. Secondary aims explored how classification accuracy 1) was affected by the timing of the sleep opportunity, 2) differed in people with versus without sleep disorders, 3) was explained by polysomnography sleep efficiency, and 4) compared to a validated consumer wearable, the Fitbit device.

## Methods

Data were utilized from 13 studies conducted at Flinders Health and Medical Research Institute: Sleep Health, from 2021-2023, where data from the WSA was collected during the sleep study. These comprised of ten studies with between 1-3 nights in the sleep laboratory for participants with suspected or diagnosed obstructive sleep apnea (OSA), insomnia, co-morbid insomnia and OSA (COMISA), cardiovascular disease, or general sleep complaint; two studies with single-night laboratory visits with healthy volunteers; and one study with two 8-day laboratory visits comprised of one nighttime sleep followed by five daytime sleep opportunities. Across all research studies, 416 sleep recordings (224 nighttime and 192 daytime recordings) with simultaneous polysomnography and WSA recordings were available for analysis. Data were time-matched based on the clock-times in the polysomnography and WSA recordings. Subsets of data were used to address secondary aims. Specifically, only data from healthy participants were available for the time of recording comparisons (given that only one study collected data from daytime sleep opportunities), only nighttime recordings were available for the healthy versus sleep disorder comparisons, and only one study of healthy participants had data available from both the WSA and the Fitbit to enable device comparisons. Detailed study information can be found in Table S1.

### Equipment

#### Polysomnography

Polysomnography was collected during all sleep opportunities. Polysomnography setups were conducted in accordance with the standard 10-20 electroencephalography electrode placement system, using Compumedics Grael 4K PSG:EEG devices (Compumedics, Victoria, Australia). Sleep studies were independently scored using Profusion Compumedics software (v 4.0) according to standardized American Academy of Sleep Medicine polysomnography scoring criteria [16]. Polysomnography sleep stages were extracted as wake, rapid eye movement (REM) sleep, stage 1 sleep (N1), stage 2 sleep (N2), and stage 3 sleep (N3).

#### Withings Sleep Analyzer

The WSA is an under-mattress device that uses a pneumatic sensor to detect changes in pressure in an air-bladder relative to atmospheric pressure (i.e., ballistography). The device uses this information to infer movement, from which respiration, heart rate and sleep stages are estimated via proprietary algorithms. The WSA was placed under the mattress, level with the chest of the sleeping individual. WSA-derived total sleep time, sleep efficiency, and wake after sleep onset has been validated, compared to polysomnography [8, 9, 11, 17], but extensive performance evaluation of sleep characteristics and sleep staging accuracy is lacking.

#### Fitbit Charge 4

The Fitbit Charge 4 is a wrist-worn device that contains a tri-axial accelerometer to track movement (i.e., actigraphy) and a photoplethysmography sensor to estimate heart rate. This information is used to infer sleep stages and wake. Fitbit devices were placed on each participant’s non-dominant wrist, with appropriate band-sized chosen to ensure proper fit. The Charge 4 model has been validated against polysomnography [18], and earlier Fitbit models similarly show reasonable accuracy to detect sleep compared to polysomnography. Individual validation studies show such devices typically overestimate total sleep time by 30-60 minutes and sleep efficiency by 5-10%, and underestimate wake after sleep onset by 20-60 minutes [19–21]. Meta-analyses suggest that newer Fitbit devices may not significantly differ from polysomnography in total sleep time, sleep efficiency, and wake after sleep onset [22], yet the latest validated devices still show poor specificity (e.g., 62% of wake correctly identified [18]). Accordingly, Fitbit devices are generally at least on-par with highly validated research-grade actiwatch devices [22, 23].

### Statistical Analysis

WSA performance was evaluated based on recommended guidelines [24, 25]. Device data (WSA and Fitbit) were extracted via custom software at www.snapi.space, developed in Python (v3.11). Polysomnography data were extracted as European Data Format files using Python, and time-matched to WSA and Fitbit data to provide concurrent epoch-by-epoch data. As the WSA device only provides sleep stage classification data 60 second intervals, Fitbit and polysomnography data were converted from 30s to 60s epochs. Where combined epochs differed, wake was scored if present in either, otherwise the first value was used [26]. Lights on and off were derived from polysomnography sleep reports to determine sleep opportunities. Where device data started or ended within these limits, preceding and trailing epochs were designated as wake.

Total sleep time (TST) was calculated as the sum of sleep epochs within the sleep opportunity. Sleep efficiency (SE) was calculated as the sum of sleep epochs divided by the total number of epochs within the sleep opportunity. Sleep onset latency (SOL) was calculated as the sum of wake epochs before the first sleep epoch, within the sleep opportunity. Wake after sleep onset (WASO) was calculated as the sum of wake epochs between the first and last sleep epoch within the sleep opportunity. Wake and sleep classification performance was determined for each sleep recording as accuracy (proportion of correctly scored epochs), sensitivity (proportion of polysomnography-derived sleep epochs that the WSA correctly scored as sleep), and specificity (proportion of polysomnography-derived wake epochs that the WSA correctly scored as wake), compared to polysomnography. Additionally, four-stage (’wake’, ’light’, ’deep’, ’REM’) sleep classification from the WSA was compared to polysomnography sleep stages, where polysomnography N2 and N3 sleep epochs were combined as ’deep’ sleep.

Linear mixed model (LMM) analyses examined 1) performance (WSA vs polysomnography) by time of recording (daytime vs nighttime) in healthy participants, 2) performance (WSA vs polysomnography) by sleep disorder status (healthy sleep vs sleep disorder), and 3) performance by consumer sleep tracker ([WSA vs polysomnography] vs [Fitbit vs polysomnography]). LMM analyses were performed using the lme4 package [27] (R v4.2.2) to examine how sleep estimates (TST, SOL, WASO, SE) and WSA performance (accuracy, sensitivity, and specificity) were affected by these three fixed effects. Participant ID was entered as random effects in all models. Marginal R^2^ was calculated to estimate variance explained by fixed effects. Secondary analysis were conducted using mixed models adjusted for age, BMI and polysomnography-derived sleep efficiency to examine potential confounders in the relationship between sleep opportunity timing, sleep disorder status, and WSA performance.

Bland-Altman plots were used to examine bias and limits of agreement (LOAs), calculated as ± 1.96 the standard deviation of mean differences and their 95% confidence limits, between polysomnography and device-derived estimates of TST, SE, WASO and SOL for each recording. Proportional bias was also calculated and tested, where significant bias indicated that the mean difference between device and polysomnography increased or decreased as a function of the size of measurement [28].

For participants with more than one recording, the coefficient of variation (CV; SD divided by mean) for accuracy, sensitivity, and specificity were calculated for each individual. To examine differences in performance variability across sleep disorder statuses and sleep opportunity timings, group differences in mean CV and the distribution of CVs were compared using linear regression and Levene tests, respectively.

Finally, in the subset where both Fitbit and WSA devices were used, data-loss was quantified and compared using a paired-samples *t*-test. Post-hoc comparisons with Bonferroni corrections were conducted where main effects were significant. All data are reported as mean (SD) unless otherwise specified. *p<*.05 was considered statistically significant.

## Results

Final data included 416 recordings from 183 participants, collected across 13 sleep research studies at Flinders Health and Medical Research Institute: Sleep Health. Participant demographics can be found in Table 1.

**Table 1:**
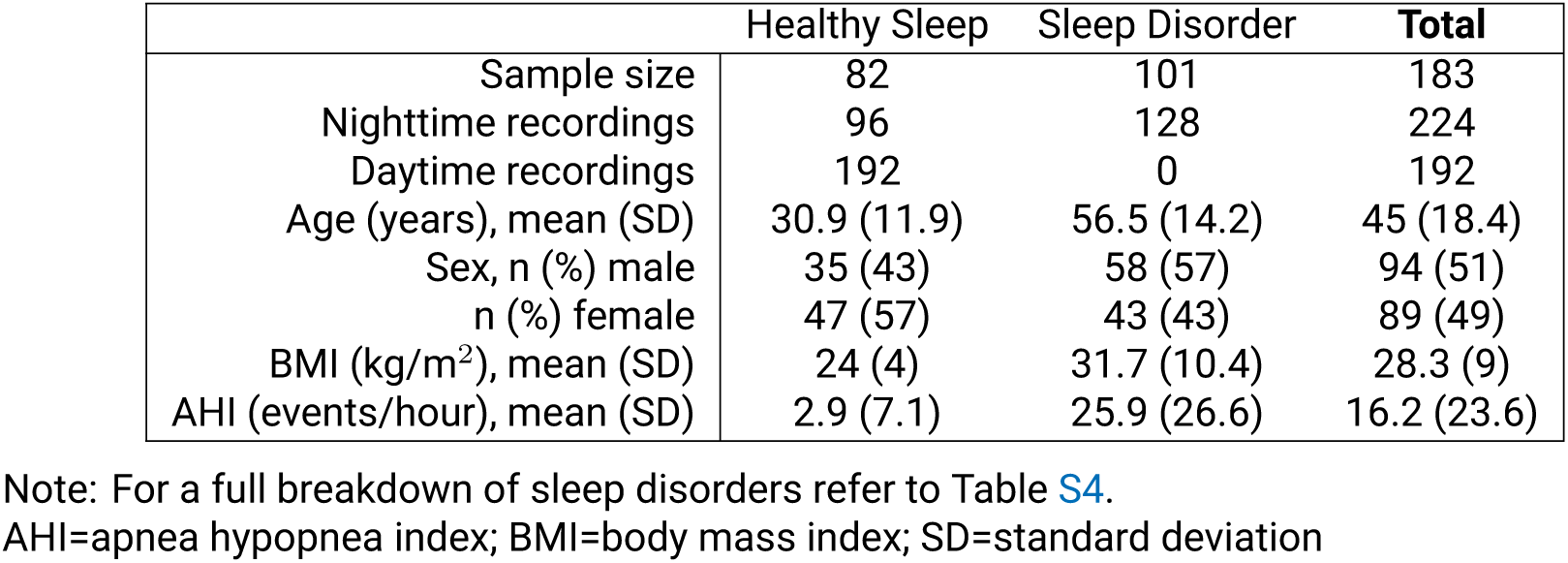
Participant demographics.

### Accuracy, Sensitivity and Specificity

The WSA had high overall accuracy and sensitivity but moderately poor specificity, compared to polysomnography, seen in Table 2. LMMs showed significantly lower mean (standard error [SEM]) accuracy of 7% (±1.4%), *F*(272)=25.2, *p<*.001, marginal R^2^=0.08, and specificity of 19.7% (±2.5%), *F*(277.2)=64.2, *p<*.001, marginal R^2^=0.18, during daytime sleep opportunities compared to nighttime sleep opportunities. There was no significant difference in sensitivity (*p>*.05). LMMs also showed significantly higher accuracy of 7.2% (±1.7%), *F*(181)=18.5, *p<*.001, marginal R^2^=0.08, and sensitivity of 4.4% (±1.5%), *F*(190.9)=8.6, *p*=.004, marginal R^2^=0.04, for healthy sleepers compared to those with a diagnosed or suspected sleep disorder.

**Table 2:** Epoch-by-epoch performance of Withings Sleep Analyzer versus polysomnography.

|  | Accuracy % | Sensitivity % | Specificity % |
| --- | --- | --- | --- |
| **Healthy Sleeps** |  |  |  |
| Nighttime | 88.7 (7.8) | 97.2 (3.3) | 46.7 (19.9) |
| Daytime | 81.8 (12) | 96.5 (6.6) | 26 (20.2) |
| **Sleep Disorders** |  |  |  |
| Nighttime | 80.6 (13.3) | 91.4 (12.6) | 46 (26.3) |
| **Overall** | 83 (12) | 95.1 (8.8) | 36.9 (24.3) |

Note: There were no data available for daytime sleep opportunities in patients with sleep disorders.

Fully adjusted models (Table 3) show that the group effect of sleep disorder status on accuracy was fully explained by age, BMI and polysomnography-derived sleep efficiency. The effect on specificity was partially explained by age. The group effect of nighttime versus daytime sleep opportunity on accuracy was partially explained by age and sleep efficiency. The effect on specificity was partially explained by sleep efficiency.

**Table 3:** Epoch-by-epoch performance of Withings Sleep Analyzer versus polysomnography.

|  | Accuracy % |  | Sensitivity % |  | Specificity % |  |
| --- | --- | --- | --- | --- | --- | --- |
| Predictor | Estimate | p | Estimate | p | Estimate | p |
| **Sleep Disorder Status** |  |  |  |  |  |  |
| Sleep Disorder | 1.95 | .271 | 4.38 | **.031** | -9.73 | **.046** |
| Age | -0.10 | **.042** | -0.01 | .844 | -0.41 | **.002** |
| Body Mass Index | -0.18 | **.018** | -0.12 | .142 | -0.12 | .550 |
| Sleep Efficiency | 0.49 | **<.001** | -0.08 | .0121 | 0.02 | .896 |
| **Sleep Opportunity Timing** |  |  |  |  |  |  |
| Daytime | -5.50 | **<.001** | -0.86 | .252 | -20.95 | **<.001** |
| Age | -0.16 | **.002** | -0.00 | .941 | -0.31 | .071 |
| Body mass index | 0.12 | .328 | 0.09 | .385 | 0.06 | .896 |
| Sleep Efficiency | 0.56 | **<.001** | 0.02 | .458 | -0.20 | **.020** |

### Sleep Characteristics

Sleep characteristics as measured by polysomnography and estimated by the WSA are found in Table 4, including subsets of data for secondary analyses. Overall, the WSA significantly overestimated TST, SE, SOL, and significantly underestimated WASO, compared to polysomnography. This was also reflected in sleep stages, where light, deep and REM sleep are typically underestimated, and wake is overestimated.

**Table 4:** Sleep characteristics of Withings Sleep Analyzer versus polysomnography during nighttime and daytime recordings.

|  | PSG | WSA | Difference (WSA - PSG) |
| --- | --- | --- | --- |
| **Overall** |  |  |  |
| Total Sleep Time | 413.1 (84.4) | 460.6 (75.3) | 47.5 (81.2)* |
| Sleep Efficiency (%) | 79.3 (14) | 88.5 (11.5) | 9.2 (14.8)* |
| Sleep Onset Latency | 17.9 (30.6) | 24.2 (29.7) | 6.4 (26.1)* |
| Wake After Sleep Onset | 90.1 (71.8) | 36.2 (59.6) | -53.9 (77.9)* |
| Wake | 108 (76.2) | 60.4 (67.3) | -47.6 (81.2)* |
| Light Sleep | 214.2 (63.3) | 238 (67.2) | 23.8 (86.4)* |
| Deep Sleep | 113.7 (44.7) | 119.4 (52.4) | 5.7 (60)* |
| REM Sleep | 84.7 (35) | 102.7 (49.2) | 18 (45.4)* |
| **Healthy Sleep - Nighttime** |  |  |  |
| Total Sleep Time | 436 (81) | 472 (70.1) | 36 (45.9)* |
| Sleep Efficiency (%) | 82.3 (12) | 89.2 (9.4) | 7 (8.6)* |
| Sleep Onset Latency | 30 (38.9) | 34.1 (39) | 4.1 (20.6) |
| Wake After Sleep Onset | 64.3 (56) | 24.1 (37.8) | -40.2 (42.5)* |
| Wake | 94.3 (68.9) | 58.3 (56.8) | -36 (45.9)* |
| Light Sleep | 253.5 (57) | 236.1 (67.2) | -17.4 (68.8) |
| Deep Sleep | 98.1 (35.8) | 129.8 (48) | 31.8 (51.2)* |
| REM Sleep | 84.2 (34) | 105.9 (38.4) | 21.7 (30.9)* |
| **Healthy Sleep - Daytime** |  |  |  |
| Total Sleep Time | 420.6 (84.5) | 486.8 (58.4) | 66.2 (79)* |
| Sleep Efficiency (%) | 78.9 (14.5) | 91.5 (9.4) | 12.7 (13.6)* |
| Sleep Onset Latency | 5.3 (5.1) | 13.5 (11.2) | 8.3 (10)* |
| Wake After Sleep Onset | 107.4 (78.2) | 32.9 (61.4) | -74.5 (79.2)* |
| Wake | 112.7 (78.4) | 46.3 (62.5) | -66.4 (78.9)* |
| Light Sleep | 192.3 (50.1) | 239.9 (60.2) | 47.6 (77.1)* |
| Deep Sleep | 132.1 (34.2) | 125.2 (38.3) | -6.9 (46.3) |
| REM Sleep | 95 (30.6) | 120.7 (45.3) | 25.7 (47.1)* |
| **Sleep Disorder - Nighttime** |  |  |  |
| Total Sleep Time | 384.6 (79.5) | 412.7 (79.2) | 28.1 (97.9)* |
| Sleep Efficiency (%) | 77.9 (14.6) | 83.5 (14.1) | 5.7 (18.8)* |
| Sleep Onset Latency | 27.6 (38) | 32.9 (35.2) | 5.2 (41.8) |
| Wake After Sleep Onset | 83.5 (65.5) | 50.2 (67.4) | -33.4 (88.4)* |
| Wake | 111.2 (77.3) | 83.1 (75.5) | -28.1 (97.9)* |
| Light Sleep | 217.4 (70.7) | 236.5 (77.1) | 19.2 (98.6) |
| Deep Sleep | 97.7 (53.6) | 102.9 (68.1) | 5.2 (76.4) |
| REM Sleep | 69.5 (36.5) | 73.2 (48.4) | 3.8 (48.7) |

Note: Data reflects mean (SD) and units are minutes unless otherwise indicated $p < .05$ in mixed model analyses between Withings Sleep Analyzer and polysomnography REM=rapid eye movement; SD=standard deviation

**Table 5:**
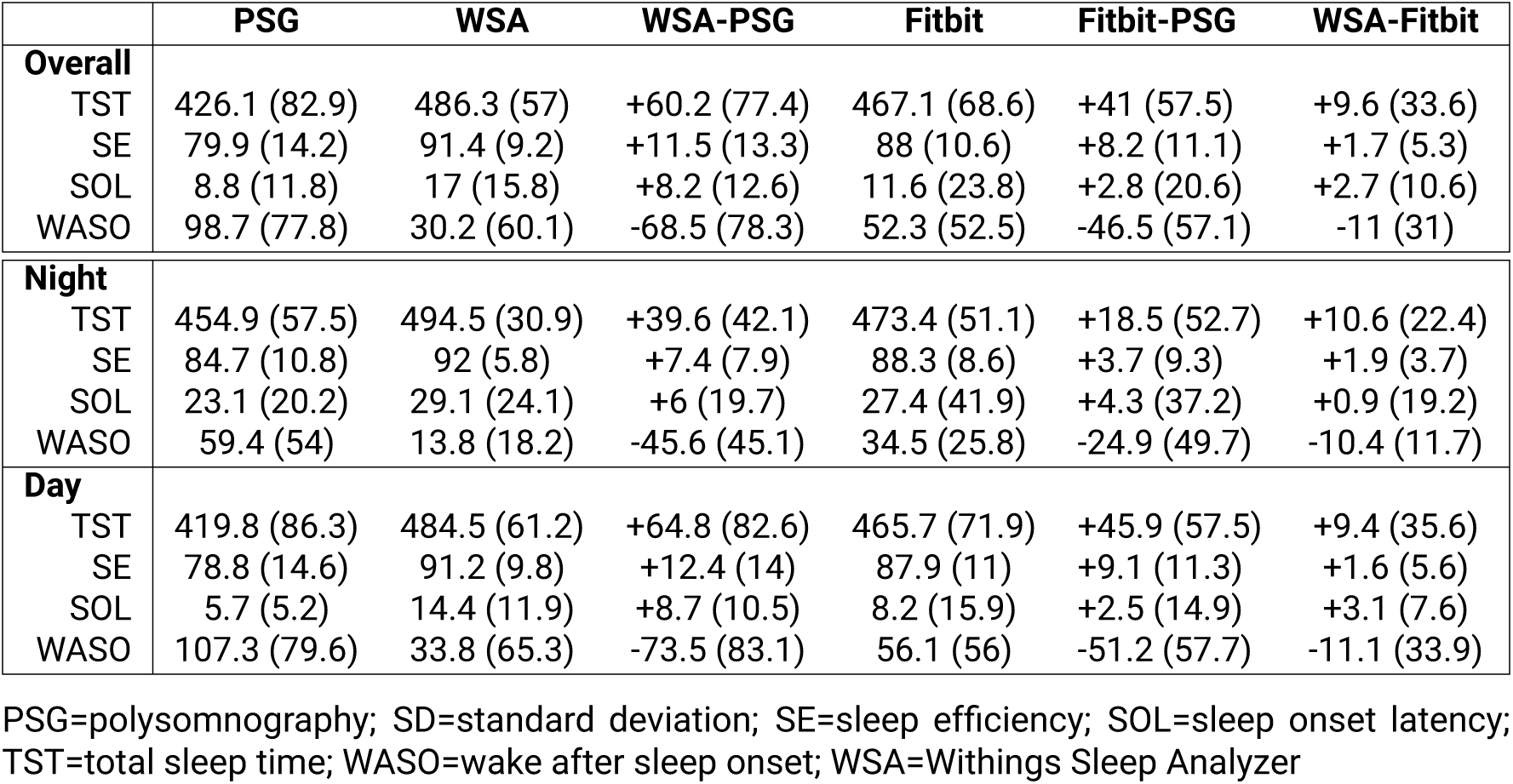
Sleep characteristics of Withings Sleep Analyzer and Fitbit versus polysomnography during nighttime and daytime recordings.

Participants had lower TST, SE, SOL and higher WASO in daytime sleep opportunities compared to nighttime sleep opportunities, and healthy participants had higher TST, SE, and lower WASO compared to those with a diagnosed or suspected sleep disorder. There were significant device by sleep opportunity timing interactions for all but SOL and REM sleep duration, where metrics were over or underestimated to a greater degree during daytime sleep opportunities. There were also significant device by sleep disorder status for light, deep, and REM sleep duration, where light sleep was overestimated and deep and REM sleep was underestimated to a greater degree in individuals with a sleep disorder, compared to healthy individuals. A detailed breakdown of interactions can be found in Supplementary Table S4.

Bland-Altman plots in Figure 1 highlight the significant increase in mean bias for TST and SE estimates, and decrease for WASO estimates, during daytime versus nighttime sleep opportunities. Furthermore, these plots show the significant proportional bias that was evident in daytime and nighttime estimated TST, SOL, and WASO, as well as daytime SE. Bland-Altman plots of healthy sleepers compared to those with a diagnosed or suspected sleep disorder did not show notable differences, as found in supplementary Figure .5.

**Figure 1:**
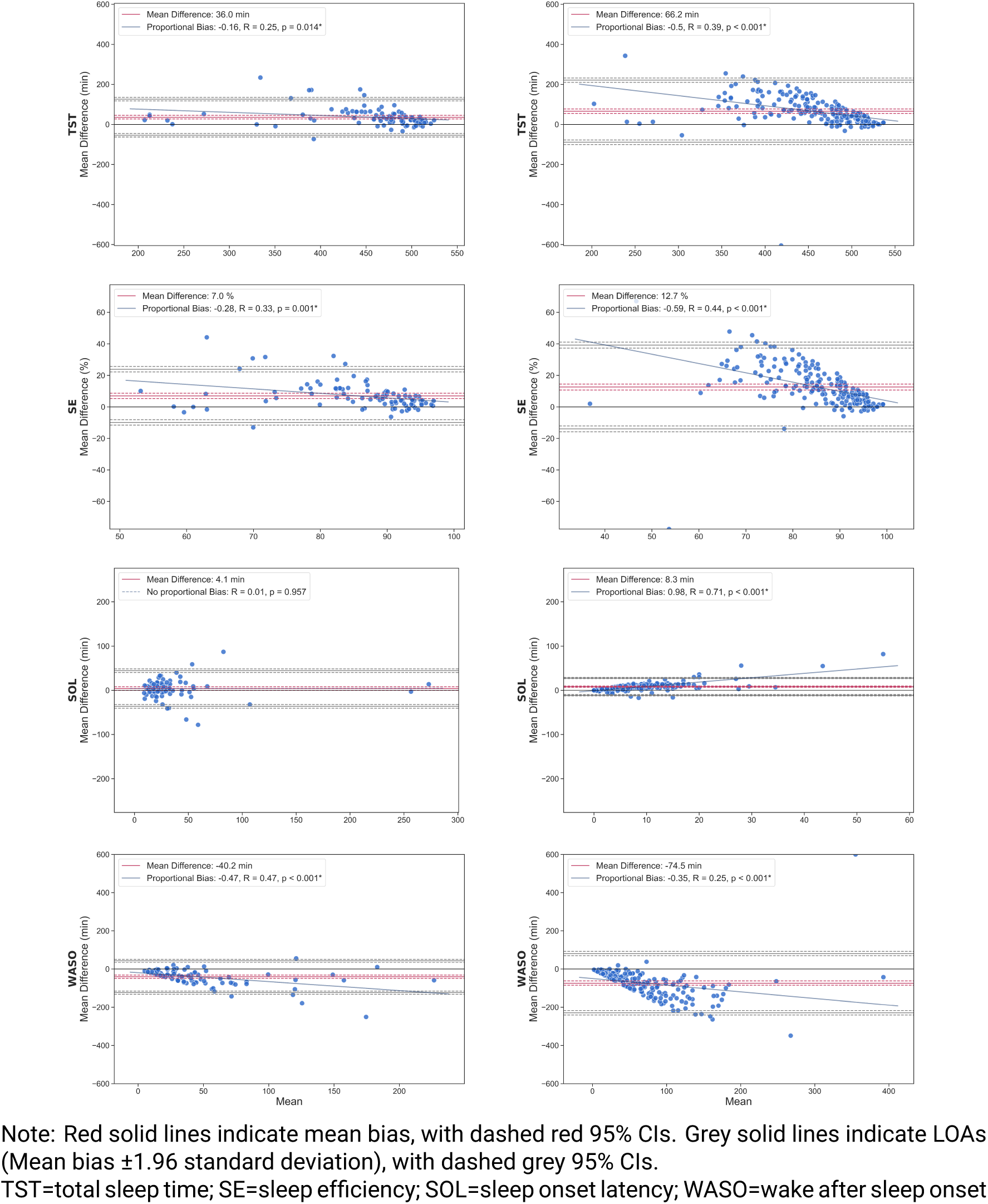
Bland-Altman plots of TST, SE, SOL, and WASO during nighttime (left) and daytime (right) sleep opportunities.

### Sleep Stages

Confusion matrices in Figure 2 demonstrate that during nighttime sleep opportunities with healthy sleepers, the WSA accurately classified 50% of wake and 91% of sleep epochs. For daytime sleep opportunities, the WSA accurately classified 27% of wake and 90% of sleep. Sensitivity for sleep stage classification ranged from 62-75% during nighttime sleep opportunities, and 60-69% during daytime sleep opportunities. For individuals with a diagnosed or suspected sleep disorder, the WSA accurately classified 43% of wake and 78% of sleep, with sleep stage accuracy ranging from 45-59%.

**Figure 2:**
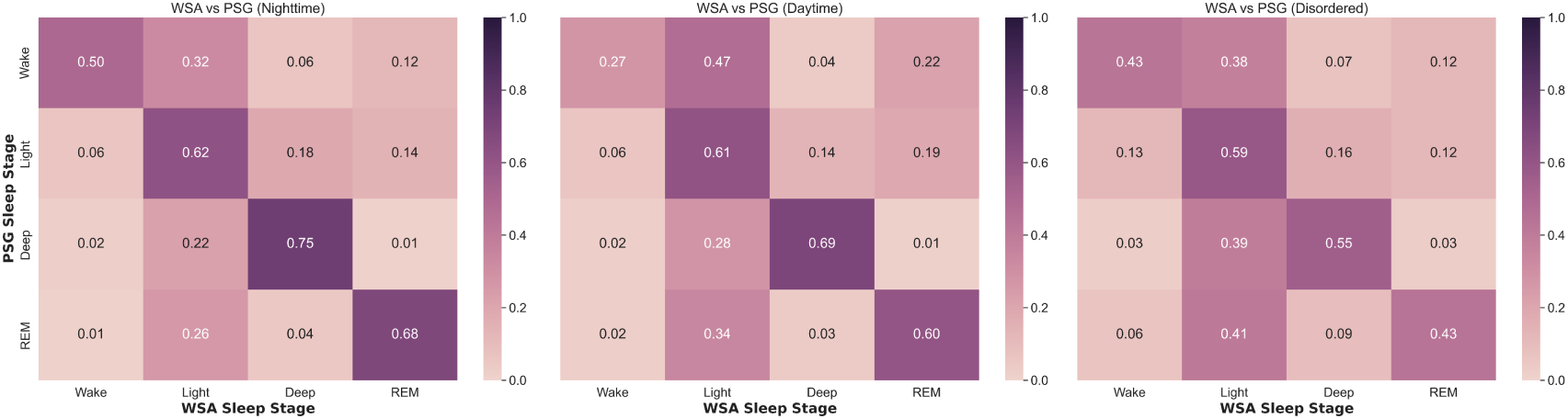
Confusion matrices showing Withings Sleep Analyzer versus polysomnography four-stage classification for healthy sleepers during nighttime and daytime recordings, and people with a sleep disorder during nighttime recordings.

### WSA versus Fitbit accuracy

Data from a subset of 22 healthy participants (nighttime recordings=35, daytime=160, mean[SD] age=31.3[12.4], 10 male, 12 female) were used to compare sleep estimation performance of the WSA and Fitbit versus polysomnography. Figure 3 shows the sleep-stage classification confusion matrices between devices for both nighttime and daytime sleep opportunities. There was a significant interaction effect of device by timing of sleep opportunity for accuracy, *F*(363.5)=6.5, *p*=.01, marginal R^2^=0.08, and specificity, *F*(365.3)=4.3, *p*=.04, marginal R^2^=0.17, whereby the Fitbit showed significantly higher accuracy (5% ±1%) and specificity (19.1% ±2.2%) than the WSA during daytime sleep opportunities, but did not significantly differ during nighttime sleep opportunities. In addition, the WSA had significantly poorer accuracy (8.7% ±1.7%) and specificity (15.7% ±3.6%) during daytime sleep opportunities, compared to night, while the Fitbit did not. This effect is evident in the confusion matrices, where daytime sleep opportunities saw a larger reduction in wake classification accuracy for the WSA (43% to 28%) compared to the Fitbit (53% to 47%). The interaction effect of device by timing of sleep opportunity was also significant, *p*=.006, but post-hoc analyses revealed no significant comparisons.

**Figure 3:**
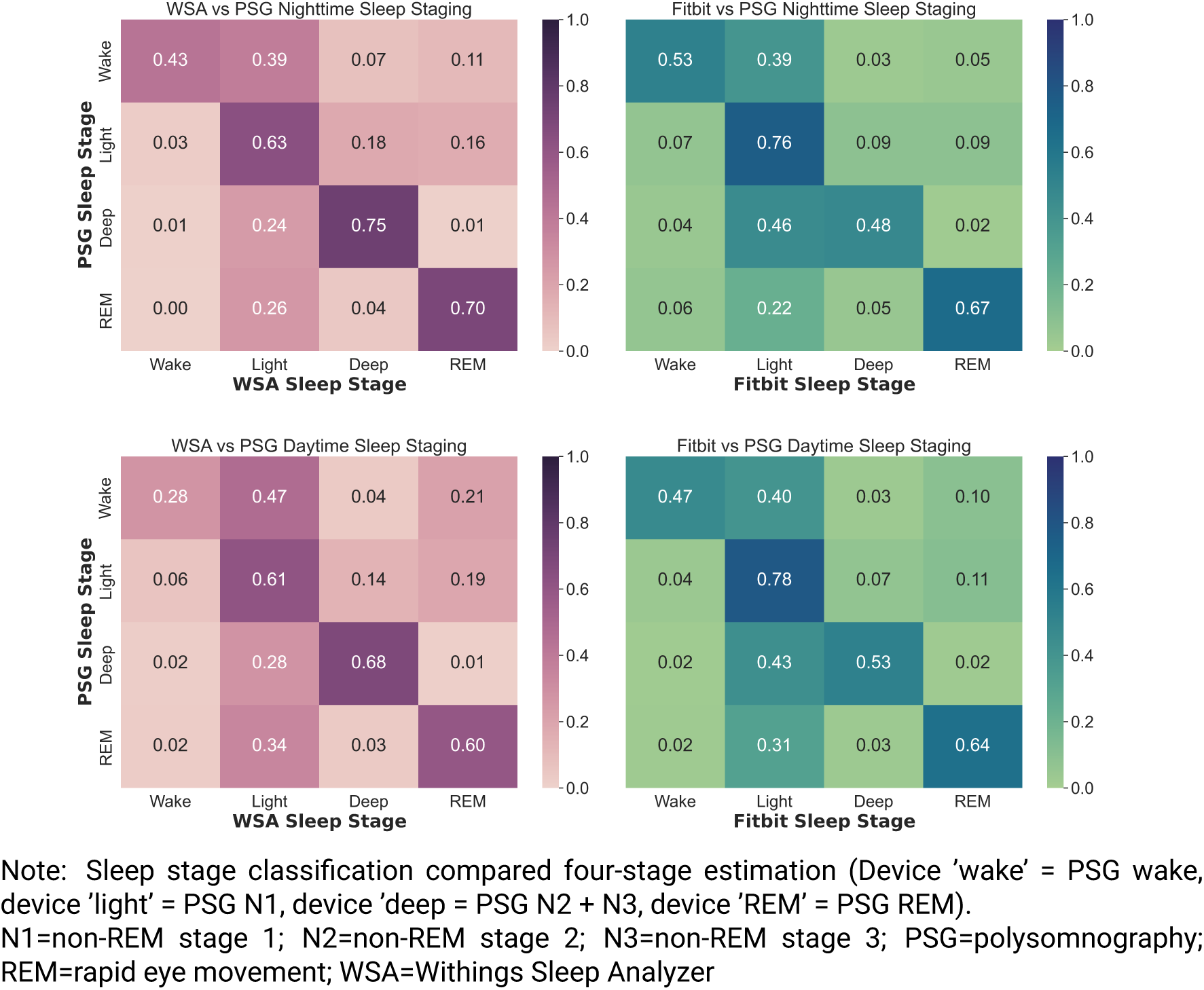
Confusion matrices showing four-stage classification, compared to polysomnography, for Withings Sleep Analyzer versus Fitbit.

Comparisons were conducted between WSA and Fitbit estimations of TST, SE and WASO, with mean (SD) values seen in Figure 5. Main effects of sleep opportunity timing and device were significant for all sleep outcomes (*ps<*.05), but there were no significant interactions between timing of sleep opportunity and device. Post-hoc analyses revealed that both the WSA and Fitbit overestimated TST and SE, and underestimated WASO, while only the WSA significantly overestimated SOL (*ps<*.05). Post-hoc analyses also showed that the Fitbit was significantly closer than the WSA to polysomnography in estimates of TST by 20 (±8.1) minutes, *p*=.04, SE by 3.5% (±1.3%), *p*=.03, and WASO by 21.5 (±7.5) minutes, *p*=.01.

### Data Loss

Within the subset where both devices were used (n=25, 248 possible recordings), a total of 17 recordings (mean [SD] = 0.68 [0.9] per participant) were lost with the WSA compared to 53 (2.12 [3.38] per participant) with the Fitbit. The paired-samples *t*-test confirmed that this was a significant difference, *t*(24) = 2.42, *p* = .02, *d* = 0.58. WSA data loss occurred entirely due to mats being unintentionally left unpowered from prior sleep studies. Fitbit data loss occurred due to improper charging (both user and mechanical error), syncing errors, and improper wear.

### Multi-night Performance

For participants with more than one recording (n=49, 282 nights), variability in accuracy, sensitivity, and specificity is demonstrated in Figure 4. Mixed models showed significantly greater variability (represented as the mean coefficient of variation) in accuracy, 2.33±2.59%, *p* = .03, and specificity, 6.03±6.85%, *p* < .001, for daytime compared to nighttime sleep opportunities. Sensitivity was more variable in individuals with a sleep disorder, 2.42±3.02%, *p* = .02. The Levene’s test also indicated that the distribution of sensitivity variability differed, 6.12, *p* = .02, seen in Figure 4 as the wider distribution of variability for individuals with disordered sleep. No other significant differences in distribution were found.

**Figure 4:**
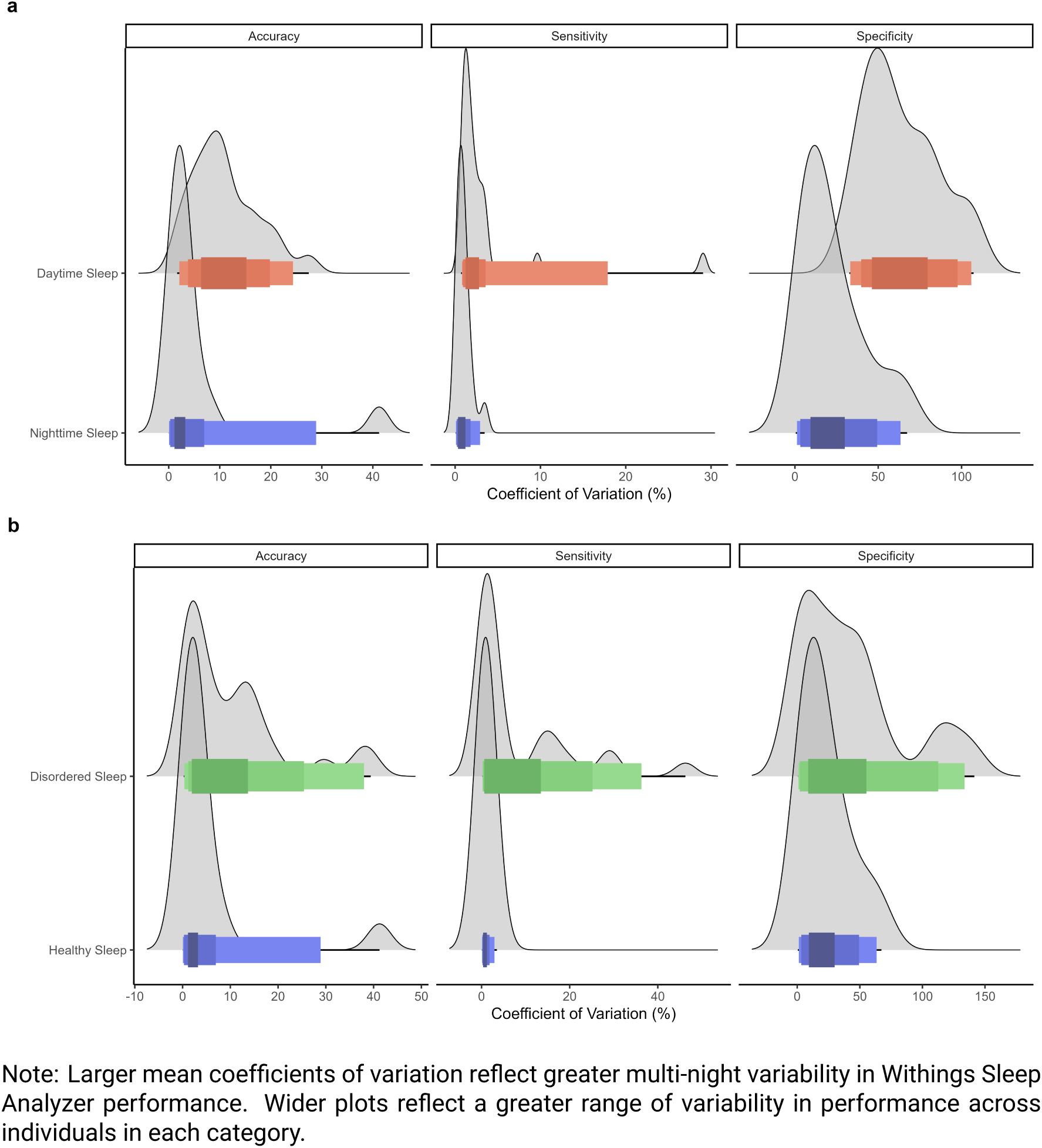
Withings Sleep Analyzer versus polysomnography variability in accuracy for a) daytime versus nighttime sleep opportunities, and b) individuals with healthy versus disordered sleep.

## Discussion

This study comprehensively evaluated the performance of the WSA to estimate sleep and wake compared to polysomnography. Appropriate use of such consumer sleep technology requires adequate performance evaluation [24]. Here, we completed one of the largest independent performance evaluations of a consumer sleep tracker to date. Results indicate that the WSA was accurate compared to polysomnography to classify sleep and wake states, with moderate-to-poor accuracy for sleep staging, that was comparable to existing wearable devices [8, 29]. The WSA systematically overestimated TST, SE and SOL, and underestimated WASO, compared to polysomnography. Accuracy and specificity was comparable to existing devices during nighttime sleep opportunities with healthy sleepers, but poorer during daytime sleep opportunities and poorer to estimate sleep in people with sleep disorders. These effects were partially explained by lower polysomnography-derived sleep efficiency during these instances. Overall, the WSA demonstrates relatively minimal bias in TST and SE estimates that is comparable to other validated consumer sleep tracking devices [8, 22], but performance was worse for sleep periods with more wakefulness.

These data are consistent with previous studies in which movement-based sleep trackers often misclassify motionless wake as sleep [30]. However, few studies have investigated circumstances where poorer than normal sleep is likely to impact reliable sleep estimation. Compared to one study with actigraphy devices [31], the WSA performs relatively poorly for reliably detecting wake during daytime recordings, with 72% of wake misclassified as sleep. However, in the subset of participants with a diagnosed or suspected sleep disorder, the lower accuracy was driven by a reduction in sensitivity (i.e., relatively more sleep being misclassified as wake). This may be due to the especially high misclassification of light sleep by the WSA as wake or deeper sleep, and the relatively high proportion of such sleep in people with a sleep disorder (OSA in particular) [32–34]. This suggests that the WSA, and likely other sleep tracking devices [18, 35–37], are less accurate to classify sleep and wake in individuals with a sleep disorder. Whether these devices are accurate enough for a given purpose is difficult to determine from the accuracy data alone. Performance evaluation, together with investigation into the potential utility of such sleep estimations, is required to make this judgement. We have shown that the WSA has utility to estimate cognitive performance under simulated shift work conditions [38], and other studies have found associations between wearable device estimations of sleep and cognitive fatigue or associated outcomes[39, 40]. Thus, further work is needed to test how the proportion of sleep misclassification observed in the current study may impact the device utility in clinical, occupational, and related contexts.

This study also showed that that the Fitbit had more reliable sleep classification accuracy than the WSA. Specifically, the Fitbit showed higher accuracy and specificity (more accurate wake classification) during daytime sleep opportunities. This was further evidenced in sleep characteristics, where the Fitbit overestimated TST by around 20 minutes less, overestimated SE by about 4% less, and underestimated WASO by around 20 minutes less than the WSA. This may be due to limitations in the modality of the two devices, where relatively motionless wake could be interpreted more accurately by a wrist-worn device than an under-mattress sensor. It may be possible that, given perceived restriction to movement imposed by polysomnography equipment, participants were less inclined to change position or move during wake. This effect would likely be more prominent in chest movement, as the WSA primarily estimates, than in wrist movements measured by the Fitbit. Other differences between device algorithms may also account for the differences between WSA and Fitbit accuracy, particularly in the incorporation of heart rate and breathing signals. Overall, the Fitbit device was more accurate at classifying sleep than the WSA, particularly during daytime sleep opportunities, but further comparisons are warranted to examine whether this is maintained in a naturalistic environment.

There are additional considerations that should be noted when critically evaluating sleep tracking devices. The WSA, unlike wrist-worn devices, does not need charging or manual synchronization of data. As such, in the study used for the sub-sample device comparison, we found a data-loss rate of 7% (compared to 21% data loss with the Fitbit). As opposed to the laboratory environment with technicians ensuring suitable device setup and suitable use to protect data fidelity, data loss would be expected to be higher in a naturalistic environment. This greater data loss may be higher for the Fitbit device than the WSA, given that, once set up, the WSA does not require user input or effort to continue recording whereas wrist-worn devices must be charges, worn correctly, and routinely synchronized for lossless data capture. Additionally, the WSA has been found to be highly accurate at bed occupancy timing (i.e., in/out of bed times) and duration compared to polysomnography with video [10], which wearable devices are typically less accurate at detecting in home environments [22, 41, 42]. Thus, the WSA has practical advantages over other forms of sleep tracking devices that should be taken into consideration when selecting a device for use, alongside its accuracy for sleep/wake detection.

Although there are strengths to the large sample size, there are still limitations to this study. Some of the included research studies with individuals with a diagnosed or suspected sleep disorder had pre-sleep events that may have impacted sleep, such as drug vs. placebo interventions and respiratory testing. Given that sleep efficiency partially explained WSA performance compared to polysomnography (by about 30-35%) the direct effects of such covariates on sleep quality likely account for some variability in WSA classification performance. However, the extensive and heterogeneous sample provides opportunity to evaluate performance with the expected variability in real-world use (e.g., variable continuout positive airway pressure use, acute sedative use, noise disruptions, etc.). It should also be considered that this study was conducted using data from entirely in-laboratory sleep research protocols, and performance in a naturalistic setting may differ. Finally, there were insufficient numbers to compare device accuracy between individual sleep disorders, and this should be elucidated in further work.

### Conclusions

This study extensively compared the WSA to polysomnography in a large sample of healthy individuals and people with a diagnosed or suspected sleep disorder during nighttime and daytime sleep opportunities to provide novel insights into performance characteristics. Overall, the WSA was accurate at sleep and wake detection during nighttime recordings compared to other consumer sleep trackers but was less accurate at wake classification during daytime recordings. The WSA was also less accurate at sleep classification in people with a suspected or diagnosed sleep disorder. The WSA overestimated TST, SE, SOL, and underestimated WASO to comparable levels seen with other consumer sleep trackers [8, 29]. Lower polysomnography-derived sleep efficiency was associated with worse WSA wake classifications, but this only partially explained the effect of wake on classification accuracy. In a subset of participants, the Fitbit was highly comparable to the WSA during nighttime recordings, but consistent with better wake classification performance, was more accurate during daytime recordings. Ultimately, the WSA may be suitable to estimate sleep and wake in a variety of naturalistic environments, and the final choice should depend on the performance, logistics, and pragmatic needs of the clinical practice or research study. The current study findings provide important novel data in which to inform these key decisions.

## Data Availability

The data that support the findings of this study are available from the corresponding author upon reasonable request.

## Acknowledgements

The authors thank and acknowledge the support from our colleagues at Flinders University and FHMRI: Sleep Health. The authors also thank all participants involved in this study.

