## Supplementary material for "Performance evaluation of an under-mattress sleep sensor versus polysomnography in *≥* 400 nights with healthy and unhealthy sleep": Table S1

Table S1: Detailed study breakdown.

| Study | N | Disorder | Nighttime | Daytime | Available Recordings | | |
|---|---|---|---|---|---|---|---|
| | | | | | Nighttime vs. Daytime Analysis | Healthy Sleep vs. Disordered Sleep Analysis | WSA vs Fitbit Analysis |
| 1 | 6 | OSA | 8 | - | - | 8 nights | - |
| 2 | 7 | OSA | 8 | - | - | 8 nights | - |
| 3 | 15 | COPD | 20 | - | - | 8 nights | - |
| 4 | 41 | None | 41 | - | 41 nights | 41 nights | - |
| 5 | 25 | None | 39 | 192 | 39 nighttime, 192 daytime | 39 nighttime | 35 nighttime, 160 daytime |
| 6 | 4 | OSA | 7 | - | - | 7 nights | - |
| 7 | 3 | OSA | 3 | - | - | 3 nights | - |
| 8 | 8 | CVD + Sleep Problem | 8 | - | - | 8 nights | - |
| 9 | 28 | OSA | 33 | - | - | 28 nights | - |
| 10 | 5 | OSA | 5 | - | - | 5 nights | - |
| 11 | 24 | 38% COMISA, 29% Insomnia, 33% OSA | 35 | - | - | 24 nights | - |
| 12 | 1 | OSA | 1 | - | - | 1 nights | - |
| 13 | 16 | None | 16 | - | 16 | 16 nights | - |
| Total | 184 | | 226 | 192 | 289 nighttime, 192 daytime | 226 nights | 35, 160 daytime |

| Study | Protocol |
|---|---|
| 1 | Single-night sleep studies with 1-3 repeats |
| 2 | Single-night sleep studies with 1-3 repeats |
| 3 | Single-night sleep studies with 1-3 repeats |
| 4 | Single-night sleep studies |
| 5 | 2 x 8-day lab stays with 1 nighttime sleep followed by 5 daytime sleeps |
| 6 | Single-night sleep studies with 1-3 repeats |
| 7 | Single-night sleep studies |
| 8 | Single-night sleep studies |
| 9 | Single-night sleep studies with 1-3 repeats |
| 10 | Single-night sleep studies |
| 11 | Single-night sleep studies with 1-3 repeats |
| 12 | Single-night sleep studies |
| 13 | Single-night sleep studies |

Note: Final Withings Sleep Analyzer versus Fitbit comparison used fewer recordings than available due to Fitbit data loss.
COMISA=Comorbid Insomnia and Sleep Apnea; COPD=Chronic Obstructive Pulmonary disease; CVD=Cardiovascular Disease; OSA=Obstructive Sleep Apnea; WSA=Withings Sleep Analyzer
