## Supplementary material for "Performance evaluation of an under-mattress sleep sensor versus polysomnography in *≥* 400 nights with healthy and unhealthy sleep": Table S2

Table S2: Model outcomes of Withings Sleep Analyzer versus polysomnography estimates between sleep disorder statuses and sleep opportunity timings.

| **Sleep Opportunity Timing** | | | | | | | | |
|---|---|---|---|---|---|---|---|---|
| | Total Sleep Time | | Sleep Efficiency | | WASO | | SOL | |
| *Predictor* | *Estimate* | *p* | *Estimate* | *p* | *Estimate* | *p* | *Estimate* | *p* |
| Device | 36.04 | <0.001 | 6.97 | <0.001 | -40.16 | <0.001 | 4.11 | 0.019 |
| Daytime | -29.39 | 0.001 | -4.54 | 0.002 | 44.67 | <0.001 | -18.20 | <0.001 |
| Device*Daytime | 30.18 | 0.005 | 5.68 | 0.002 | -34.33 | 0.001 | 4.15 | 0.054 |
| | Wake | | Light Sleep | | Deep Sleep | | REM Sleep | |
| *Predictor* | *Estimate* | *p* | *Estimate* | *p* | *Estimate* | *p* | *Estimate* | *p* |
| Device | -36.04 | <0.001 | -17.44 | 0.014 | 31.75 | <0.001 | 21.73 | <0.001 |
| Daytime | 24.65 | 0.004 | -60.45 | <0.001 | 25.85 | <0.001 | 7.92 | 0.094 |
| Device*Daytime | -30.34 | 0.004 | 65.03 | <0.001 | -38.63 | <0.001 | 3.94 | 0.509 |

| **Sleep Disorder Status** | | | | | | | | |
|---|---|---|---|---|---|---|---|---|
| | Total Sleep Time | | Sleep Efficiency | | WASO | | SOL | |
| *Predictor* | *Estimate* | *p* | *Estimate* | *p* | *Estimate* | *p* | *Estimate* | *p* |
| Device | 28.12 | <0.001 | 5.67 | <0.001 | -33.36 | <0.001 | 5.24 | 0.123 |
| Disordered | 48.73 | <0.001 | 4.12 | 0.025 | -18.83 | 0.024 | 3.53 | 0.526 |
| Device*Disordered | 7.92 | 0.470 | 1.30 | 0.522 | -6.80 | 0.484 | -1.13 | 0.828 |
| | Wake | | Light Sleep | | Deep Sleep | | REM Sleep | |
| *Predictor* | *Estimate* | *p* | *Estimate* | *p* | *Estimate* | *p* | *Estimate* | *p* |
| Device | -28.12 | <0.001 | 19.16 | 0.008 | 5.18 | 0.368 | 3.77 | 0.339 |
| Disordered | -15.38 | 0.130 | 38.98 | <0.001 | -3.60 | 0.634 | 13.88 | 0.016 |
| Device*Disordered | -7.92 | 0.461 | -36.60 | 0.001 | 26.57 | 0.003 | 17.96 | 0.003 |

Note: Device estimates reflect PSG-WSA. Daytime/Disordered reflects estimate relative to nighttime/healthy.

PSG=polysomnography; REM=rapid eye movement; SOL=sleep onset latency; WASO=wake after sleep onset; WSA=Withings Sleep Analyzer.
