## Supplementary material for "Performance evaluation of an under-mattress sleep sensor versus polysomnography in *≥* 400 nights with healthy and unhealthy sleep": Figure S1

Figure S1: Bland-Altman plots of TST, SE, SOL, and WASO for healthy participants (left) and participants with suspected or diagnosed sleep disorders (right).

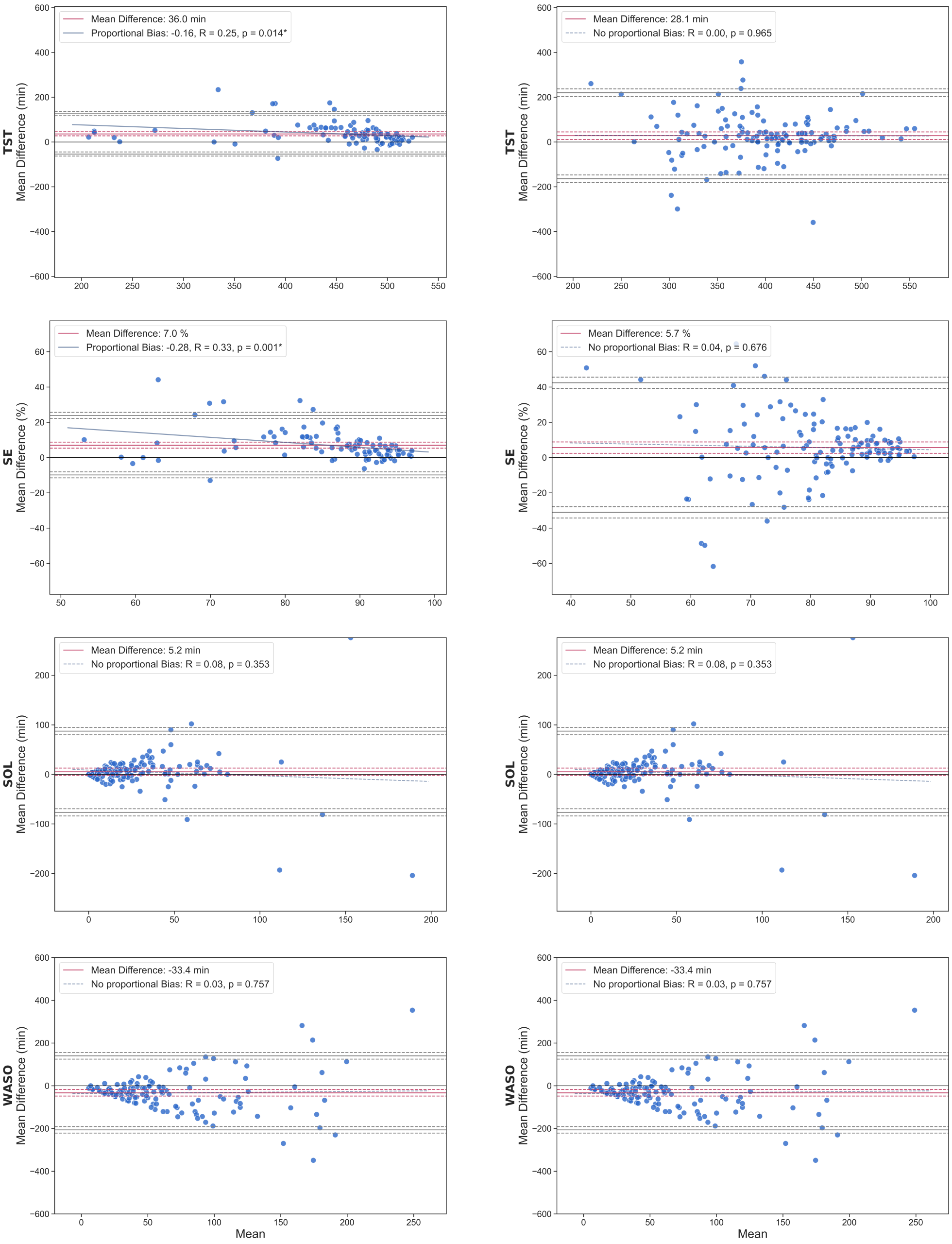


**Note:** Red solid lines indicate mean bias, with dashed red 95% CIs. Grey solid lines indicate limits of agreement (Mean bias ±1.96 standard deviation), with dashed grey 95% CIs. Blue solid lines indicate significant ($p<.05$) proportional bias. Blue dashed lines indicate non-significant bias. TST=total sleep time; SE=sleep efficiency; SOL=sleep onset latency; WASO=wake after sleep onset
